# High proportion of Ugandans with pre-pandemic SARS-CoV-2 cross-reactive CD4+ and CD8+ T-cell responses

**DOI:** 10.1101/2023.01.16.23284626

**Authors:** Annemarie Namuniina, Enoch S Muyanja, Victoria M Biribawa, Brenda A Okech, Aloysious Ssemaganda, Matt A Price, Nancy Hills, Ann Nanteza, Bernard Ssentalo Bagaya, Daniela Weiskopf, Catherine Riou, Steven J Reynolds, Ronald M Galwango, Andrew D Redd

## Abstract

The estimated mortality rate of the SARS-CoV-2 pandemic varied greatly around the world with multiple countries in East, Central, and West Africa having significantly lower rates of COVID-19 related fatalities than many resource-rich nations with significantly earlier wide-spread access to life-saving vaccines. One possible reason for this lower mortality could be the presence of pre-existing cross-reactive immunological responses in these areas of the world. To explore this hypothesis, stored peripheral blood mononuclear cells (PBMC) from Ugandans collected from 2015-2017 prior to the COVID-19 pandemic (n=29) and from hospitalized Ugandan COVID-19 patients (n=3) were examined using flow-cytometry for the presence of pre-existing SARS-CoV-2 cross-reactive CD4+ and CD8+ T-cell populations using four T-cell epitope mega pools. Of pre-pandemic participants, 89.7% (26/29) had either CD4+ or CD8+, or both, SARS-CoV-2 specific T-cell responses. Specifically, CD4+ T-cell reactivity (72.4%) and CD8+ T-cell reactivity (65.5%) were relatively similar, and 13 participants (44.8%) had both types of cross-reactive types of T-cells present. There were no significant differences in response by sex in the population. The rates of cross-reactive T-cell populations in these Ugandans is higher than previous estimates from resource-rich countries like the United States (20-50% reactivity). It is unclear what role, if any, this cross-reactivity played in decreasing COVID-19 related mortality in Uganda and other African countries, but does suggest that a better understanding of global pre-existing immunological cross-reactivity could be an informative data of epidemiological intelligence moving forward.

## Background

The severe acute respiratory syndrome coronavirus-2 (SARS-CoV-2) causes Coronavirus disease 2019 (COVID-19), a communicable respiratory disease with symptoms ranging from asymptomatic to severe acute respiratory distress in humans. Disease presentation is likely affected by a complex array of factors including host genetics, pre-existing immune status, sex, age, and nutritional status [1].

COVID-19 mortality was significantly higher in the western world compared to Eastern, Central, and West Africa, and this was true despite the fact that broad access to life-saving COVID-19 vaccines was inequitably delayed for much of Africa. The global excess mortality rate between January 2020 to December 2021 has been estimated to be 120.3 deaths per 100,000 (10^5^) people, but this rate varied widely from >500 per 10^5^ people in some countries in Eastern Europe to no COVID-related deaths in countries with total isolation strategies [2]. While excess mortality was influenced by many factors, the trends suggest that countries in West, Central, and East Africa were generally protected from the worst COVID-19-related mortality. For example, during this period the estimated increase in mortality in Uganda was 93.5 per 10^5^ people, whereas the rate was 179.3 per 10^5^ people in the United States, 227.4 per 10^5^ people in Italy, and a shocking 647.3 per 10^5^ people in Bulgaria [2]. While there are certainly many social, demographic, and equity factors that influence these estimates, it is likely that levels of underlying immunological cross-reactivity to SARS-CoV-2 could also affect mortality.

A pre-existing immune response to other circulating human common cold coronaviruses (hCCCoV) is thought to decrease the severity of COVID-19. In one study, the presence of immunoglobulin G (IgG) antibodies against the SARS-CoV-2 receptor binding domain (RBD) and pre-existing common cold coronaviruses were tested in hospitalized patients, and those with high IgG levels had milder disease compared to those with low or no detectable IgG [3]. A study in the United States found that individuals who had a known hCCCoV infection the year before the SARS-CoV-2 pandemic had significantly lower rates of mortality and severe disease compared to individuals without previous infection the year before [4]. Furthermore, a recent study examined pre-existing anti-SARS-COV-2 humoral responses among populations in France and several African countries and found that pre-pandemic African samples were approximately ten times more likely to be serologically reactive to SARS-CoV-2 compared to the French participants [5].

In addition to antibody responses, individuals with a high level of pre-existing memory CD4+ T-cells that are cross-reactive with SARS-CoV-2 may mount a faster and stronger immune response, thereby limiting disease severity [6]. It has been proposed that SARS-CoV-2-specific T-cells in non-exposed individuals originate from memory T-cells derived from previous hCCCoV exposure, which is common in the human population [7,8]. Several studies have observed that 20-50% of people who had not been exposed to SARS-CoV-2 had presence of cross-reactive CD4+ and/or CD8+ T-cells, a phenomenon thought to occur due to sequence similarity between immunodominant coronavirus epitopes [9–13]. In the United States, Grifoni et. al. observed cross-reactivity in up to 50% of donor blood samples obtained between 2015 and 2018, prior to the appearance of SARS-CoV-2 in the human population [11]. T-cell cross-reactivity was greatest against proteins other than the SARS-CoV-2 spike protein, but T-cell cross-reactivity against spike was also observed. The majority of SARS-CoV-2 T-cell reactivity was associated with CD4+ T-cells, with a minor contribution from CD8+ T-cells. In the Netherlands CD4+ T-cell cross-reactivity against SARS-CoV-2 spike peptides was observed in 10% of unexposed individuals, while reactivity to SARS-CoV-2 non-spike peptides was seen in 20% of unexposed individuals [10]. A study in Germany found cross-reactive T-cell responses to spike peptides in 34% of SARS-CoV-2 unexposed individuals [9]. Similarly, T-cell cross-reactivity to nucleocapsid protein non-structural protein (nsp7 or nsp13) was found in 50% of individuals with no history of SARS, COVID-19, or contact with SARS or COVID-19 patients in a study carried out in Singapore [12,14].

Given the lower COVID-19 mortality noted in many parts of Africa and the expected role that pre-existing cross-reactive immunity may have on disease severity, this study aimed to investigate the presence and magnitude of SARS-CoV-2 cross-reactive T-cells in pre-pandemic Ugandans.

## Methods

### Study scope and design

Frozen peripheral blood mononuclear cells (PBMC) collected during the Simulated Vaccine Efficacy Trial (SIVET) were used for this analysis. The goal of the SIVET study was to assess whether people from Ugandan fishing communities could be enrolled, vaccinated, and retained in a simulated vaccine efficacy trial using licensed Hepatitis B and Typhoid vaccines in place of experimental vaccines (manuscript in preparation). Briefly, PBMC were collected from participants aged 18 to 49 years between 2015-2017 from one fishing community in Entebbe along the shores of Lake Victoria before the COVID-19 pandemic. 40ml of blood was drawn and PBMCs isolated by density gradient centrifugation and stored in liquid nitrogen at 10 million cells per vial. As part of the SIVET study, participants were tested for HIV, schistosomiasis, and Hepatitis B. The testing kits used for HIV were Determine, StatPak and Unigold, for schistosomiasis Kato Katz was used, and Hepatitis B testing was performed with VIDAS® HBs Ag Ultra, VIDAS® Anti-HBs Total II, and VIDAS® Anti-HBc Total II assays. Participants provided written informed consent to participate in the study, and agreed to the use of their samples in future related research studies. All samples were collected and processed >1 year before the emergence of SARS-CoV-2. For this study, we used 28 samples from participant enrollment visits, and 1 participant follow up visit, six months later.

In addition, a group of Ugandan patients hospitalized due to complications from COVID-19, were included as positive controls. 8ml of blood was collected, anonymized, and PBMC (5×10^6^) were isolated by density gradient centrifugation and stored in liquid nitrogen. No demographics data was collected from patients.

### Ethical approval

The study was approved by the National Council for science and technology (NCST) and original SiVET study was also approved by the Uganda Virus Research Institute Research and Ethics Committee (UVRI REC), GC/127/841. Participants provided written informed consent to participate in the study.

### Determination of Cell viability

10 million PBMC (10×10^6^ cells/ml) were thawed in a 37^°^C water bath for one minute. Before completion of thawing, the cells were transferred from the water bath to a 50ml sterile tube containing 10ml R10 media (complete RPMI with 10% fetal calf serum) and 20μL Dnase (20µl/10ml). The mixture was spun for at 1200 rpm (revolution per minute) for seven min at 4^°^C. The supernatant was discarded and cells were re-suspended in 1ml of R10 media and cell concentration was determined by examining 20µl (1:1) of a mixture of PBMC/Trypan blue (0.4%) on a hemocytometer.

### Activation Induced cell Marker (AIM) assay

The previously described SARS-CoV-2 epitope MegaPool (MP) preparations were used to examine for possible reactive T-cell populations [10]. A detailed description on the T-cell predictions carried is available in the following manuscript [15].

The AIM assay was used to detect the antigen specific T-cells responses after PBMC stimulation [16,17]. Briefly, the MP used targeted CD4+ T-cells with spike-specific epitopes (CD4_S; n=253) or non-spike epitopes (CD4_R, n=221). Additionally, CD8+ T-cell responses were examined using the CD8_A and CD8_B epitopes which were estimated to interact with the 12 most common HLA class I A and B alleles [10,11].

T-cell activation was determined as described previously [11]. Briefly, cells were cultured for 24 hours in 96-well U bottom plates with 1.5×10^6^ PBMC per well in the presence of SARS-CoV-2 specific MPs (1 mg/ml). A stimulation with an equimolar amount of DMSO was used as the negative control, while Phytohemagglutinin (PHA, Roche, 1 mg/ml) and the combined CD4 and CD8 Cytomegalovirus MP (CMV, 1 mg/ml) were used as positive controls, as previously described.

### Flow Cytometry

#### PBMC immune cell phenotyping

After stimulation, cells were washed in 200µl PBS at 1400rpm at 4^°^C for 2 min. For the surface stain, 1.5 ×10^6^ PBMCs were resuspended in 100 µl Magnetic-Activated Cell Sorting (MACS) buffer and stained with antibody cocktail for 30 min at 4^°^C in the dark (Supplemental Table S1). Following cell surface staining, cells were washed twice with MACS buffer. Cells were resuspended in 100 µl PBS and kept at 4^°^C before acquiring on the BD LSRII SORP flow cytometer (BD Biosciences).

### Flow cytometer gating

Following acquisition T-cell populations were interrogated as shown in the gating strategy to identify reactive CD4+ and CD8+ T-cell populations (Supplementary Figure S1). Briefly, cells were initially gated according to acquisition time to remove artifacts like air bubbles or clogs. The CD3+ cell population was then selected via Forward Scatter (FSC) and Side Scatter (SSC) and segregated according to CD3 expression to select lymphocytes. This was followed by singlet gating to remove doublets. This was further followed by gating on live cells. The live cells were divided into CD4+ and CD8+ populations and examined for activation by the Antigen Induced Markers (AIM) CD137+, OX40+ for CD4+ T-cells and CD69+, CD137+ for CD8+ T-cells, which were both presented in percentages of total CD4+ or CD8+ T-cells. An average number of 150,000 cells was acquired. Responses for both CD4+ and CD8+ cells were examined for all four megapools.

### Data analysis

Data analysis was done using Flowjo (version 10.8, FlowJo LCC, Ashland, OR, USA), Stata (version 17.0; College Station, TX, USA), and GraphPad Prism (version 9; GraphPad Software Inc, San Diego, CA, USA). Initially, responsiveness was visually examined for all samples, and any sample with no visual reactivity for a given MP was set at 0%. For all responders, total CD4+ and CD8+ T-cell response was calculated by subtracting the percentage of activated positive cell responses after SARS-CoV-2 MP stimulation from the percentage of cell responses after DMSO stimulation. The lowest value across the four SARS-CoV-2 peptides was used if the percentage of AIM positive cell responses after DMSO stimulation was zero.

## Results

PBMC samples from our comparison group of actively hospitalized COVID-19 patients were initially analyzed for SARS-CoV-2 T-cell reactivity. Five samples were originally examined, but two were excluded for low CD4 % and poor CD4 staining, leaving only three for final analysis (Figure 1, Supplemental Table S2). All three had both CD4+ and CD8+ reactive T-cell populations, although not all patients responded to all four MPs tested (Supplemental Table S2).

**Figure 1:**
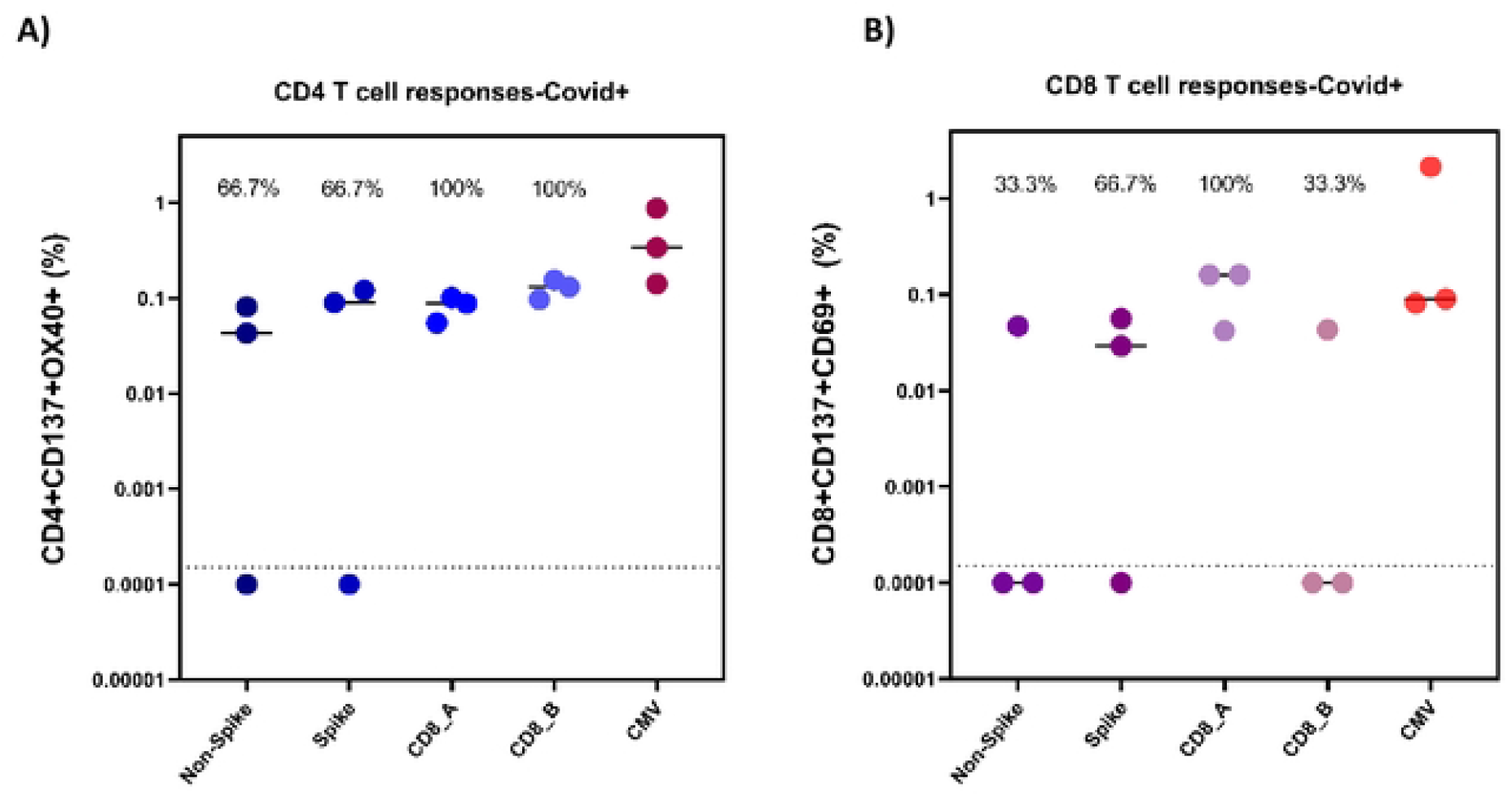
CD4+ (A) and CD8+ (B) T cell responses in COVID-19 hospitalized patients (n=3). Reactive cell percentages of total CD4+ or CD8+ T cells are shown for CD4_Non-spike, CD4_Spike, CD8_A, CD8_B, and CMV megapools for both T cellpopulations.

The pre-pandemic analysis data set (n=29 participants) included 21 males (72%) with participants being younger with a median age of 27 years (Table 1). Samples from these participants were collected between Dec 2015 and May 2017, and were from fishing communities on the shores of Lake Victoria. These communities tend to be crowded with a significant amount of migration in and out of the area throughout the year. All participants were HIV negative since the original SiVET study aimed to recruit only HIV-uninfected individuals. Out of the 29 participants, four people (13.8%) were infected with Schistosomiasis, and no cases of hepatitis infection were observed. Per the original study protocol, those participants with Hepatitis B test results suggesting pre-existing immunity to Hepatitis B (i.e., previously vaccinated before joining the study) were not vaccinated at enrolment.

**Table 1.**
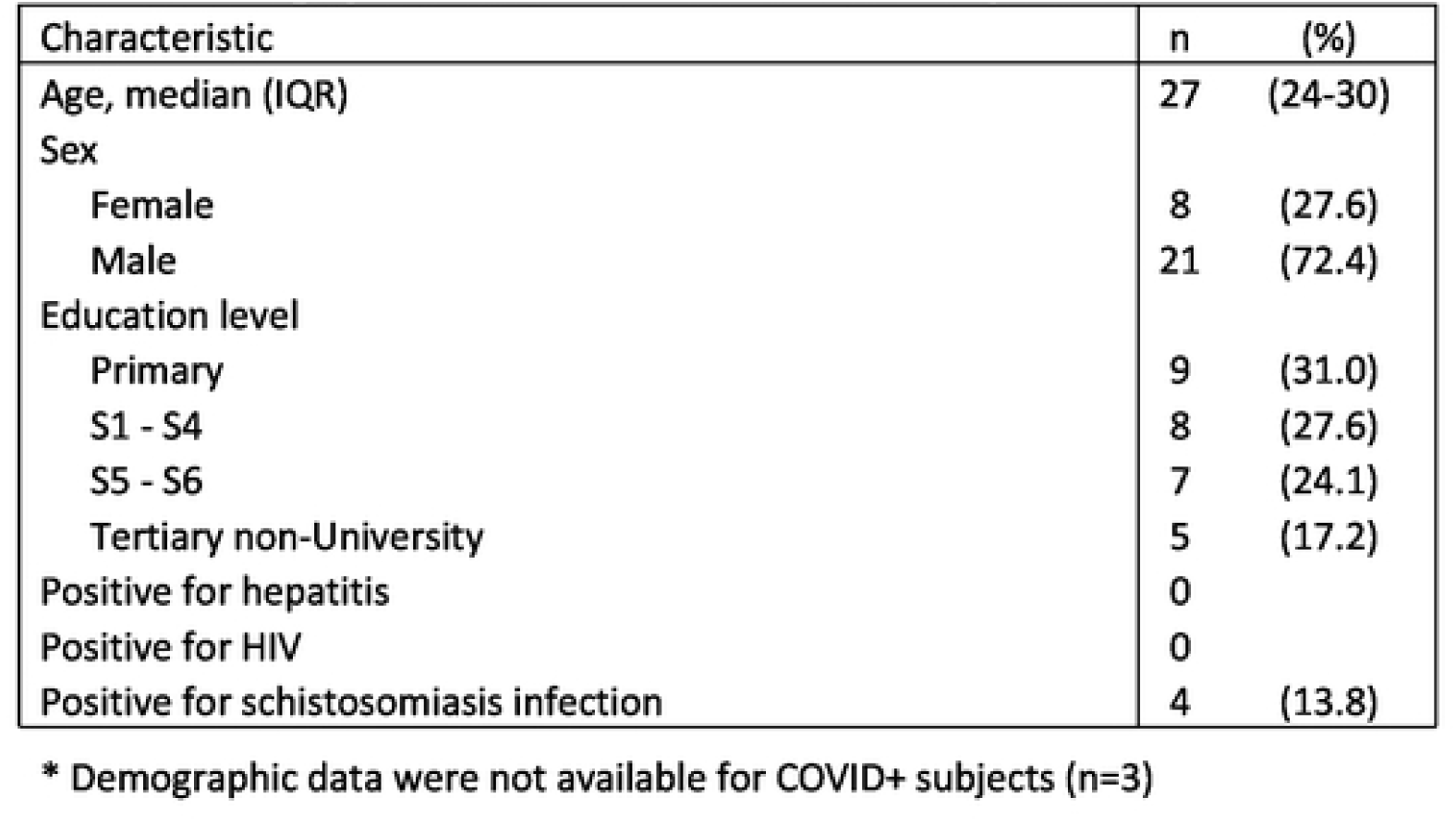
Demographic characteristics of 29 non-COVID patients*.

| Characteristic | n | (%) |
| --- | --- | --- |
| Age, median (IQR) | 27 | (24-30) |
| Sex |  |  |
| Female | 8 | (27.6) |
| Male | 21 | (72.4) |
| Education level |  |  |
| Primary | 9 | (31.0) |
| S1 - S4 | 8 | (27.6) |
| S5 - S6 | 7 | (24.1) |
| Tertiary non-University | 5 | (17.2) |
| Positive for hepatitis | 0 |  |
| Positive for HIV | 0 |  |
| Positive for schistosomiasis infection | 4 | (13.8) |

\* Demographic data were not available for COVID+ subjects (n=3)

In the pre-pandemic samples (n=29), it was found that 44.8%, 58.6%, 31.0%, and 41.4% of participants had a detectable CD4+ T-cell response to the CD4_R (non-spike), CD4_S (spike), CD8_A, and CD8_B peptide pools, respectively (Figure 2A and Table 2). Furthermore, it was revealed that 72.4% (21) of participants had CD4+ T -cell response to at least one MP, and 17.2% of individuals were responsive to all four MP tested (Table 2).

**Figure 2:**
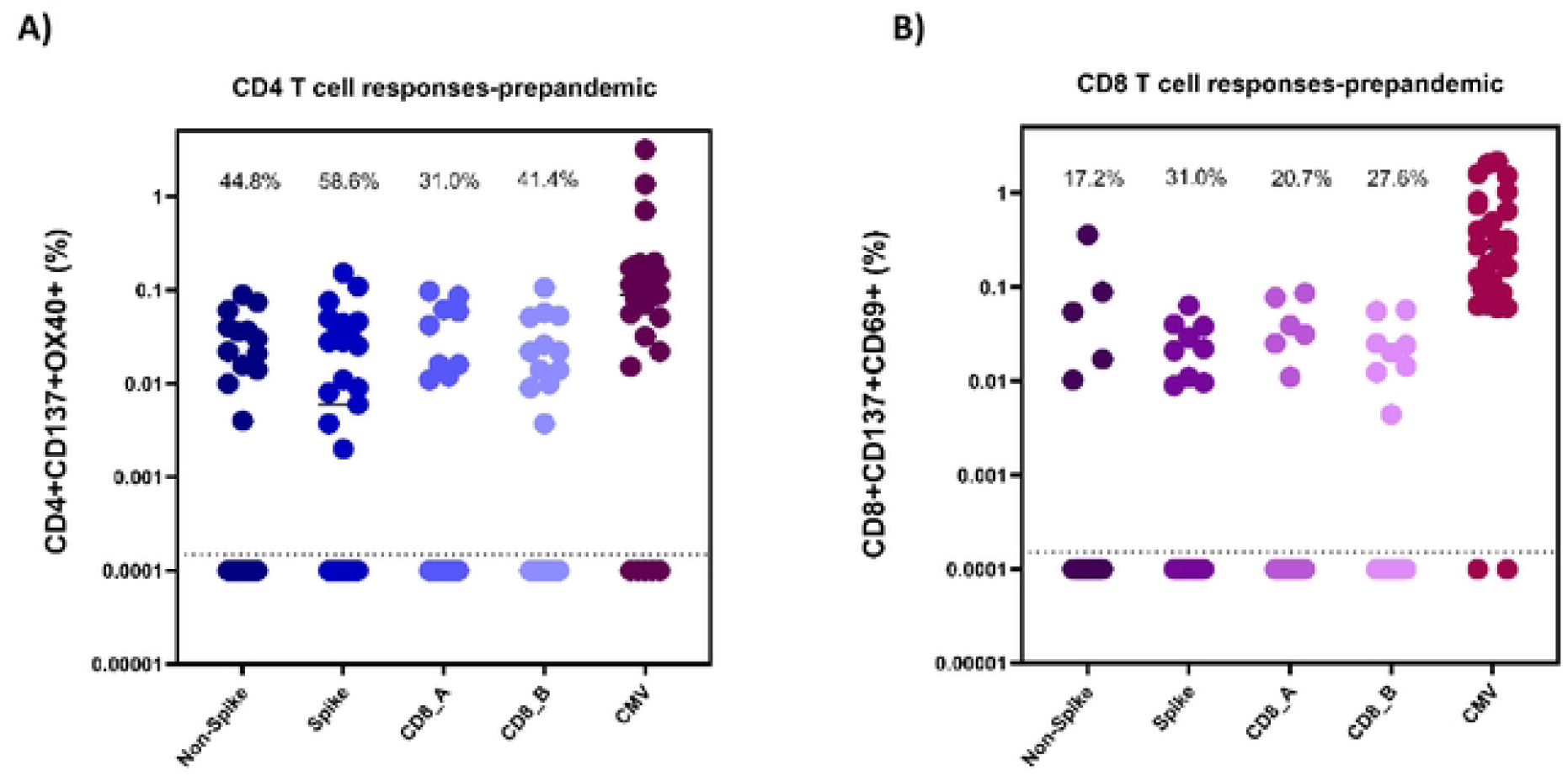
CD4+ (A) and CD8+ (8) T cell responses in pre-pandemic Ugandan PBMC samples (n=29). Reactive cell percentages of total CD4+ or CDS+ T cells are shown for CD4_Non-spike, CD4_Spike, CD8_A, CD8_B, and CMV megapools for both T cell populations.

**Table 2.** CD4+ and CD8+ T cell responses after stimulation for pre-pandemic participants.

| *Pre-pandemic cohort, n=29* | CD4+ T cell Reactivity | % CD4+ Reactivity | CD8+ T cell Reactivity | % CD8 Reactivity |
| --- | --- | --- | --- | --- |
| *CD4_Non-spike* | 13 | 44.8% | 5 | 17.2% |
| *CD4_Spike* | 17 | 58.6% | 9 | 31.0% |
| *CD8_A* | 9 | 31.0% | 6 | 20.7% |
| *CD8_B* | 12 | 41.4% | 8 | 27.6% |
| *Total Pools Reactive* |  |  |  |  |
| 0 | 8 | 27.6% | 10 | 34.5% |
| 1 | 6 | 20.7% | 9 | 31.0% |
| 2 | 5 | 17.2% | 5 | 17.2% |
| 3 | 5 | 17.2% | 3 | 10.3% |
| 4 | 5 | 17.2% | 0 | 0.0% |
|  | No Reactive T cells | CD4+ T Cell reactive only | CD8+ T cells reactive only | Both T cells reactive |
| *# of participants* | 3 | 9 | 4 | 13 |
| *% of participant* | 10.3% | 31.0% | 13.8% | 44.8% |

CD8+ T-cell reactivity was slightly lower for each MP tested with 17.2%, 31.0%, 20.7%, and 27.6% of individuals having reactivity to the CD4_R (non-spike), CD4_S (spike), CD8_A, and CD8_B peptide pools, respectively (Figure 2B and Table 2). It was found that 65.5% (19) of participants had some CD8+ T-cell response to at least one MP, and 10.3% of individuals were responsive to three of the four MP tested (Table 2). No individuals had reactive CD8+ T-cells to all four MP tested.

Taken together these data demonstrate that 89.7% (26/29) of this Ugandan population had some detectable T-cell response (either CD4+ or CD8+) pre-pandemic, and 44.8% (13/29) had both CD4+ and CD8+ reactive T-cells (Table 2). In addition, the responsiveness was similar in both males and females in this cohort (Table 3).

**Table 3.**
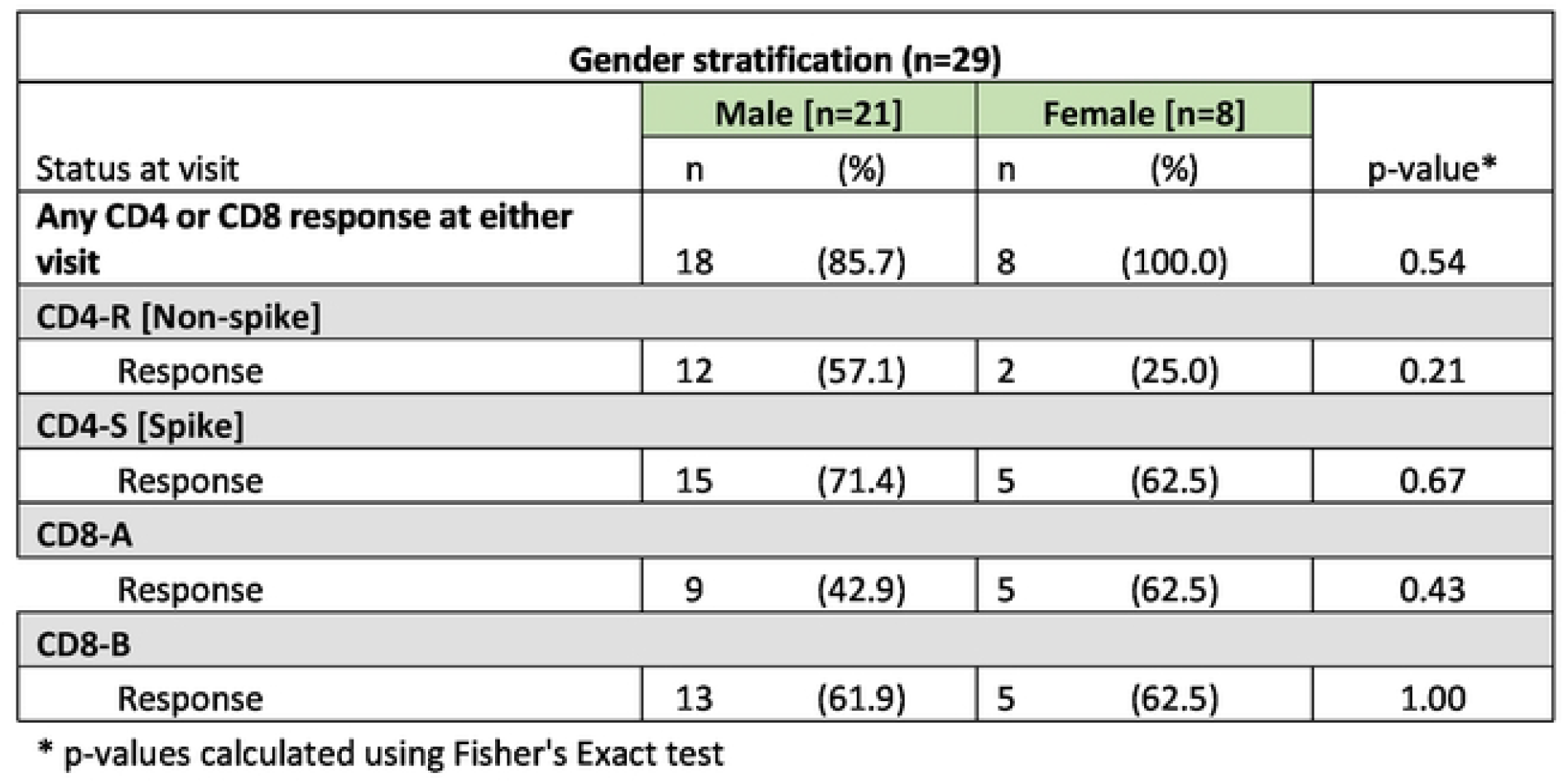
Gender and T Cell responses.

## Discussion

There is a need to investigate the cause of disproportionate COVID-19 disease severity in sub-Saharan Africa as compared to western countries. In this study, cross-reactive T-cell responses to SARS-CoV-2 were observed in 90% of adult Ugandans in samples collected between Dec 2015 and May 2017, which was well before the onset of the pandemic. It has been speculated that these cross-reactive immunological responses are due to exposure to hCCCoVs that share sequence homology and structure with SARS-CoV-2 [8,11]. T-cell derived immunity plays a critical role in our full immunological response to novel pathogens. It has been shown that participants with pre-existing CD4 T-cell reactivity were able to mount a faster spike-specific CD4 and antibody response following subsequent COVID-19 vaccination [18]. It is possible that this rapid memory T-cell response results in a more protective response in the event a person becomes exposed to SARS-CoV-2. However, it should be pointed out that some studies have suggested that the presence of cross-reactive T-cell responses may not offer protection and could cause greater disease severity in COVID-19 patients [19]. Either way, the relatively high proportion of Ugandans with cross-reactive T-cells demonstrated here suggest these pre-existing responses might have been more prevalent in some African populations compared to the western world where mortality rates were significantly higher [9–11].

It should be noted that previous cross-reactivity studies in African populations have focused on humoral responses, whereas this study focused on T-cell responses. However, other studies have shown that when compared to antibodies and CD8+ T-cells, SARS-CoV-2-specific CD4+ T-cells had the strongest association with reduced COVID-19 disease severity [20]. In addition, the absence of SARS-CoV-2-specific CD4+ T-cells was linked to severe or fatal COVID-19 infections [9]. These findings of a higher proportion of cross-reactive T-cell responses in Ugandans is supported by the finding that pre-exposure samples from Central, East, and Western Africa were more likely to have cross-reactive antibody responses to SARS-CoV-2 than comparable samples from France and USA [5,21].

This study had several limitations. The sample size of the study was relatively small due to limited sample availability and issues with low CD4+ cell percentage after cell acquisition. However, the sample size is comparable to other similar studies from resource-rich countries. Additionally, it was not possible to further characterise responses in memory T-cell subsets because the cell number and total events collected were too low to accurately measure the rare events in both COVID+ and pre-pandemic samples. Finally, previous exposure to other hCCCoVs was not examined in this study.

In summary, high levels of both SARS-CoV-2 specific CD4+ and CD8+ T-cell responses was observed in this group of Ugandans well before the COVID-19 pandemic. Further work is needed to fully elucidate the role that these cross-reactive responses may have played in the relatively low COVID-19 related mortality rates in some areas of Africa

## Data Availability

Data availability: Anonymized versions of the data presented here is available upon request and pending approval of all pertinent review committees.

## Funding information

This work was supported by NIH contract 75N93019C00065 (A.S, D.W), and in part by the Division of Intramural Research, NIAID, NIH. The authors wish to acknowledge the support from the University of California, San Francisco’s International Traineeships in AIDS Prevention Studies (ITAPS), U.S. NIMH, R25MH123256. Funding for the original SIVET was provided by IAVI. This work was made possible by generous support from the United States Agency for International Development (USAID). The full list of IAVI donors is available at www.iavi.org. The contents are the responsibility of the authors and do not necessarily reflect the views of USAID or the United States Government

## Acknowledgements

The authors would like to thank the study participants and UVRI-IAVI study staff.

## Data availability

Anonymized versions of the data presented here is available upon request and pending approval of all pertinent review committees.

## Competing interests

LJI has filed for patent protection for various aspects of T cell epitope and vaccine design work.

---

**Supplementary Table S1:**
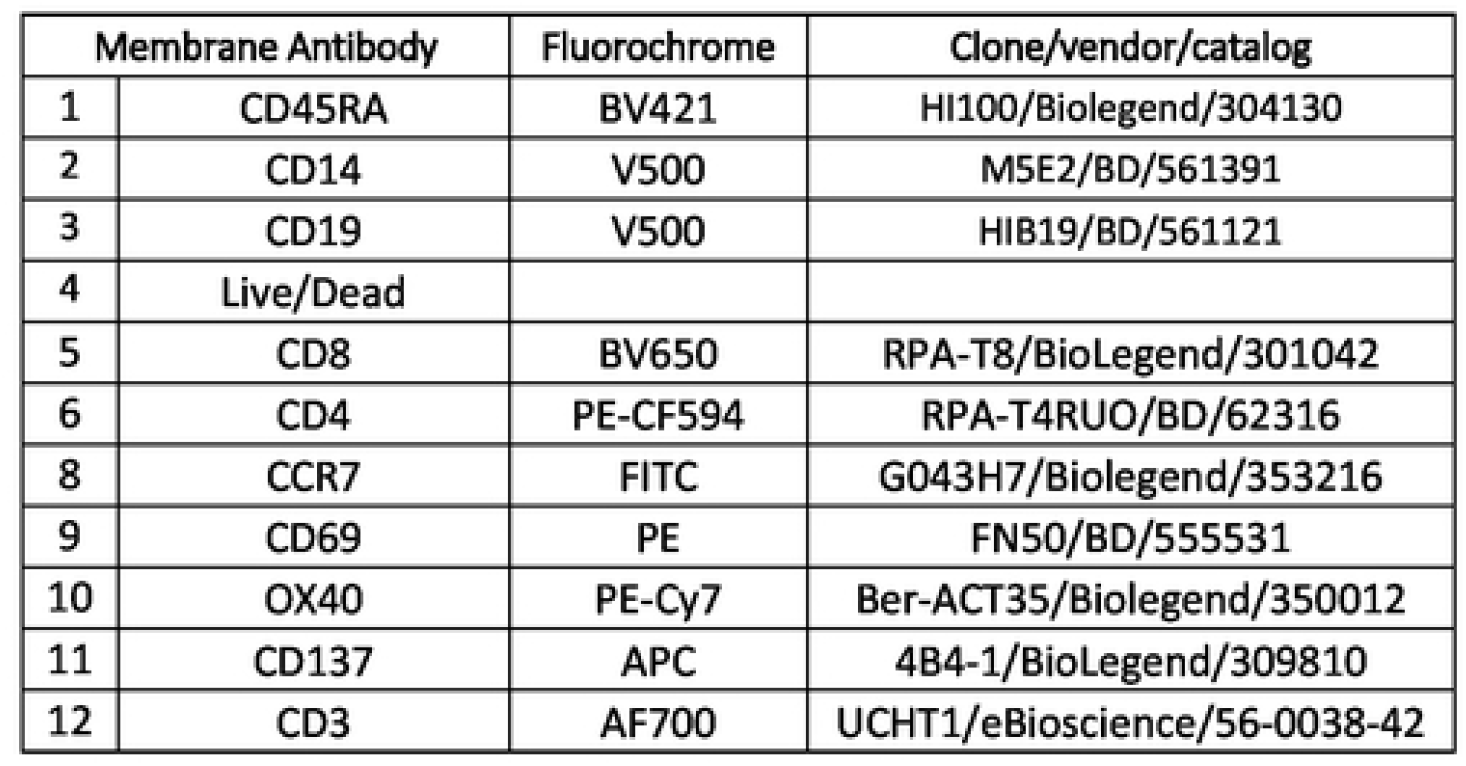
AIM antibodies used for cell staining.

**Supplementary Table S2:** Hospitalized COVID + samples

| *COVID cohort, n=3* | CD4+ T cell<br>Reactivity | % CD4+ Reactivity | CD8+ T cell<br>Reactivity | % CD8<br>Reactivity |
| --- | --- | --- | --- | --- |
| *CD4_Non-spike* | 2 | 66.7% | 1 | 33.3% |
| *CD4_Spike* | 2 | 66.7% | 2 | 66.7% |
| *CD8_A* | 3 | 100.0% | 3 | 100.0% |
| *CD8_B* | 3 | 100.0% | 1 | 33.3% |
| *Total Pools Reactive* |  |  |  |  |
| *0* | 0 | 0.0% | 0 | 0.0% |
| *1* | 0 | 0.0% | 1 | 33.3% |
| *2* | 1 | 33.3% | 1 | 33.3% |
| *3* | 0 | 0.0% | 0 | 0.0% |
| *4* | 2 | 66.7% | 1 | 33.3% |

**Supplementary Figure S1:**
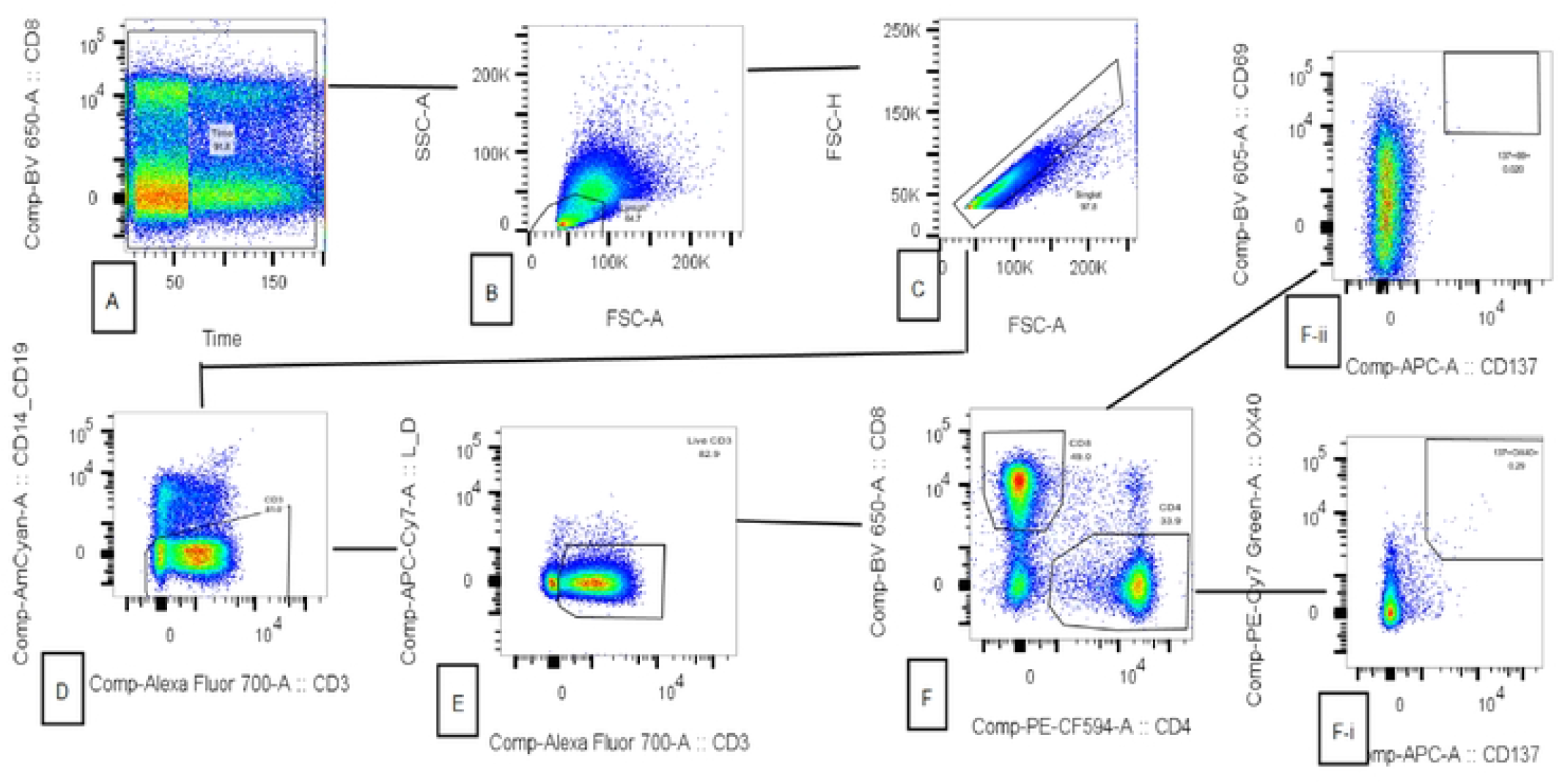
Gating strategy(A-F) for detection of SARSD-CoV-2 reactive C04+ and CD8+ cells after PBMC stimulation: Time gating was done to eliminate any artifact like air bubble **(A)**, followed by a selection of lymphocyte population **(B)**, and singlets **(C)**. Live C03+cells were selected **(D-E)**, then divided into CD4+ and CD8+ cells **(F)**. Within CD4 and CD8 subsets, antigen-specific T cells were established through the upregulation of activation-induced markers OX40, CD69, and CD137. Percentages of OX40+CD137+ double-positive cells within the CD4 gate **(F-i)** and percentage of CD69+CD137+ within the CD8 gate **(F-ii)** showing activated cells, were gated out to be used for further analysis

